# Trends in antimicrobial resistance amongst *Salmonella* Typhi in Bangladesh: a 24-year retrospective observational study (1999–2022)

**DOI:** 10.1101/2023.12.21.23300147

**Authors:** Arif M Tanmoy, Yogesh Hooda, Mohammad S I Sajib, Hafizur Rahman, Anik Sarkar, Dipu Das, Nazrul Islam, Naito Kanon, Md. Asadur Rahman, Denise O Garrett, Hubert P Endtz, Stephen P Luby, Mohammod Shahidullah, Md. Ruhul Amin, Jahangir Alam, Mohammed Hanif, Samir K Saha, Senjuti Saha

## Abstract

**Background:** Rising antimicrobial resistance (AMR) in *Salmonella* Typhi restricts typhoid treatment options, heightening concerns for pan-oral drug-resistant outbreaks. Bangladesh contemplates introducing typhoid conjugate vaccine (TCV) to address the typhoid burden and AMR. However, large-scale surveillance data on typhoid AMR in Bangladesh is scarce.

**Objective:** This study explores the AMR trends in *Salmonella* Typhi isolates from Bangladesh, drawing comparisons with antibiotic consumption to optimize antibiotic stewardship strategies for the country.

**Methods:** Our typhoid fever surveillance included two pediatric hospitals and three private clinics in Dhaka, Bangladesh, spanning 1999 to 2022. Blood cultures were performed at physicians’ discretion; cases were confirmed by microbiological culture, serological, and biochemical tests. Antimicrobial susceptibility was determined following CLSI guidelines. National antibiotic consumption data for cotrimoxazole, ciprofloxacin, and azithromycin was obtained from IQVIA-MIDAS database for comparison.

**Results:** Our 24-year surveillance, encompassing 12,435 *Salmonella* Typhi cases, revealed declining trends in first-line drugs (amoxicillin, chloramphenicol, cotrimoxazole) and multidrug resistance (MDR; 38% to 17%, 1999–2022). Cotrimoxazole consumption dropped, 0.8 to 0.1 DDD/1000/day (1999–2020). Ciprofloxacin non-susceptibility persisted (>90%) with unchanged consumption (1.1-1.3 DDD/1000/day, 2002–2020). Low ceftriaxone resistance (<1%) was observed, with rising MIC (0.03 to 0.12 mg/L, 1999–2019). Azithromycin consumption increased (0.1 to 3.8 DDD/1000/day, 1999–2020), but resistance remained ≤4%.

**Conclusion:** Our study highlights declining MDR amongst *Salmonella* Typhi in Bangladesh, thus reintroducing first-line antimicrobials could work as an empirical treatment option for typhoid fever. Our analysis provides a baseline for monitoring the impact of future interventions, including the TCV, on typhoid burden and associated AMR.

## Introduction

Antimicrobial resistance (AMR) poses a significant threat to public health, estimated to cause approximately 1.27 million deaths in 2019.^1^ If unchecked, deaths linked to AMR infections could multiply 10-fold by 2050 due to widespread antimicrobial use and a paucity of new antimicrobials.^2, 3^ In response to this threat, multiple strategies have been proposed, ranging from antibiotic stewardship to alternative antibiotic therapies. Implementing these interventions requires guidance from surveillance data, but many low- and middle-income countries (LMICs) face substantial data deficits.^1^ This study aims to provide insights into AMR trends in Bangladesh for *Salmonella enterica* serovar Typhi (Salmonella Typhi), the etiological agent of typhoid fever, over a 24-year period, from 1999 to 2022.

Typhoid fever is a pressing global public health concern, with the majority of cases occurring in South Asia.^4^ In Bangladesh, the typhoid burden is alarmingly high, especially in the capital city of Dhaka, where the estimated incidence is 913 per 100,000 person-years.^5^ While the disease is more prevalent among school-age children,^6^ those under 2 years old experience the highest bacterial load.^7^ Clinical and epidemiological data on typhoid fever have been traditionally reported from hospital-based surveillance studies, but, in many endemic countries, like Bangladesh, a significant number of typhoid patients seek care at community-based diagnostic centers or, hospital outpatient departments (OPD).^8–11^ This diverse care-seeking behavior complicates the precise estimation of the disease burden and AMR trends for typhoid fever.

Management of typhoid fever is becoming increasingly difficult with rising AMR. Multi-drug resistant (MDR, defined as the concurrent resistance to amoxicillin, chloramphenicol, and co-trimoxazole) *Salmonella* Typhi isolates emerged in the 1970s.^12, 13^ This led to ciprofloxacin as the primary treatment choice by mid-1980s, but resistance followed. By mid-2010s, over 90% of *Salmonella* Typhi in South Asia showed non-susceptibility to ciprofloxacin.^14^ The current treatments include azithromycin and third-generation cephalosporins such as ceftriaxone,^13^ but, reports of resistance to both drugs are increasing.^15–20^ The 2016 typhoid outbreak in Pakistan, caused by an extensively drug resistant (XDR; resistant to ceftriaxone, ciprofloxacin and first-line of antimicrobials) lineage,^19^ sparked global concern. Additionally, a novel *acrB*-717 gene mutation linked to azithromycin resistance,^16^ has narrowed treatment options and raised fears of a pan-oral drug-resistant (PoDR) outbreak. A recent pan-oral resistant typhoid case has been reported from Pakistan and required intravenous carbapenem and colistin treatment.^21^ This highlights the looming threat of AMR and related financial burden on patients and healthcare systems which is struggling to cope with high typhoid prevalence in LMICs.^13, 22^

To curb this high burden of typhoid fever, the Government of Bangladesh is considering the introduction of a typhoid conjugate vaccine (TCV). Phase-III trials have showed that the vaccine reduces typhoid fever by 81–85% and it has been introduced in Pakistan, Liberia, Zimbabwe, Samoa, and Nepal.^23, 24^ However, the continuous monitoring of AMR trends in the post-vaccination era is essential to ensure sustained support from policymakers. This is especially crucial for Bangladesh, which is poised to shed its Least-Developed Countries (LDC) status in 2026 and start losing financial support from Gavi for vaccines. Amidst many competing priorities, policymakers of Bangladesh will require robust surveillance data to make informed decisions about the TCV program. To this end, our study provides insights on the historical trends in AMR through a comprehensive typhoid surveillance dataset of 12,489 *Salmonella* Typhi isolates, spanning 24 years (1999-2022). It elucidates correlations between AMR and antibiotic consumption, providing a baseline to guide the design of antibiotic stewardship policies. It also provides insights into designing empirical treatment strategies for typhoid fever management and potential strategies to combat increasing AMR in Bangladesh.

## Methods and Materials

### Enteric fever surveillance

The Child Health Research Foundation (CHRF) has been conducting enteric fever surveillance in Bangladesh since 1999, with a focus on the pediatric population (<18 years of age). This surveillance is carried out at three sites in Dhaka, the capital of Bangladesh with the highest burden of typhoid fever.^6^ These sites are: (a) Bangladesh Shishu Hospital and Institute (BSHI), a 650-bed hospital that is the largest pediatric hospital in the country, (b) Dr. M R Khan Shishu Hospital & Institute of Child Health (SSFH), a 250-bed hospital that is the second largest pediatric hospital in Dhaka, and (c) three branches of the Popular Diagnostic Center (PDC), a community-based consultation and diagnostic center. All cases were classified based on whether they were treated in the outpatient facility (OPD) or admitted to the hospitals (in-patient, IPD).

All cases from PDC were classified as OPD. Blood samples were collected for microbiological culture as a part of the diagnostic service at the attending physicians’ discretion. BSHI and SSFH are part of the Global Invasive Vaccine-Preventable Bacterial Disease Surveillance Network since 2009.^25^ Details of all three sites and our enteric fever surveillance program have been described previously.^8, 26^ All data used in this study are given in Supplementary Data S1.

### Etiological confirmation and antimicrobial susceptibility testing

In all hospitals, routine blood cultures were performed using standard methods, as described earlier.^8, 27^ *Salmonella* Typhi isolates from PDC were reconfirmed at BSHI using biochemical tests (Klingler’s Iron agar, Simmons citrate agar, motility-indole, and urea agar tests) and *Salmonella* agglutinating antisera (Thermo Scientific, MA, USA). Antibiotic susceptibility tests were conducted for amoxicillin, cotrimoxazole, chloramphenicol, ciprofloxacin, ceftriaxone, and azithromycin using the Kirby-Bauer disc diffusion method (Oxoid, Thermo Scientific, MA, USA). The minimum inhibitory concentrations (MICs) for ceftriaxone and ciprofloxacin were determined for two subsets of isolates (from 1999–2019 and 1999–2016 respectively), using the broth microdilution method (Sigma-Aldrich, MO, USA).^28, 29^ All isolates collected before 2017 underwent MIC testing if recoverable on plates. Additionally, ceftriaxone MIC testing was performed on a random set of isolates from the later period (2017–2019). Zone-diameters (ZD) and MIC data were interpreted based on the guideline of Clinical and Laboratory Standards Institute (CLSI), 2020. Azithromycin-resistant (ZD: <11 mm) and near-resistant (ZD: 11-13 mm) isolates were reconfirmed using Etest strips (bioMérieux, Marcy-l’Étoile, France). The MIC_50_ for ceftriaxone and ciprofloxacin, indicating the MIC value at which 50% of tested isolates could not grow, was also calculated. Ceftriaxone MIC of two isolates (MIC >256 mg/L) were excluded from all related analyses, to mitigate the impact of outliers.

### Acquisition and analyses of antibiotic consumption data

To compare the patterns of typhoid cases, we acquired antibiotic consumption data (1999-2020) for co-trimoxazole, ciprofloxacin, and azithromycin from the IQVIA-Multinational Integrated Data Analysis System (IQVIA-MIDAS®) database. This commercial database captures retail pharmacy sales, encompassing the total antibiotic volumes sold to both retailers and hospital pharmacies by wholesalers. It also records annual sales of each antimicrobial agent for each country. We excluded antimicrobial sales data for the agricultural sector. Daily defined doses (DDD) for these antimicrobials were gathered from ATC/DDD Index 2023 (https://www.whocc.no/atc_ddd_index/). DDD represents the assumed average maintenance dose per day for its main indication in adults. Adjusting for population size, we calculated DDDs per 1000 inhabitants per day, following the WHO’s Global Antimicrobial Resistance and Use Surveillance System (GLASS).^30^ Annual population data for Bangladesh was collected from the United Nations-World Population Prospect Report 2019.^31^

### Statistical analysis

We used Pearson’s correlation coefficients to assess correlations between yearly AMR patterns and consumption of co-trimoxazole, ciprofloxacin, and azithromycin. All correlation tests were conducted in Stata v13.0. We also examined potential non-linear resistance patterns for ciprofloxacin and ceftriaxone using their MIC data, and for azithromycin with yearly resistance percentage data. The analyses for ciprofloxacin and ceftriaxone employed the generalized additive model (GAM) with the *mgcv* package in R (version R 4.2.3). The model formula was *y ∼ s(x, bs = “cs”)*, with 3,214 and 3,518 observations for ciprofloxacin and ceftriaxone, respectively. Yearly trend of azithromycin resistance was analyzed similarly in R (version R 4.2.3) using local polynomial regression.

## Results

### Resistance to the first-line antimicrobials amoxicillin, chloramphenicol, cotrimoxazole

Our typhoid fever surveillance recorded 12,435 *Salmonella* Typhi cases from 1999 to 2022 (Data S1). Of these cases, 28% (3,478/12,435) required hospitalization (IPD) at either BSHI or SSFH hospital (Figure S1). During this period, we observed a declining trend in resistance among *Salmonella* Typhi isolates to the first-line antimicrobials—amoxicillin, chloramphenicol, and cotrimoxazole. In 2002, resistance peaked at 80%, but decreased to less than 20% in recent years for all three first-line antimicrobials (Figure 1a-c). This decline was also reflected in multidrug resistance (MDR), which was 17% in 2022 (Figure 1d). The percentage of MDR isolates remained ≤26% since 2010 (average 20%; 18%–23% at 95% Confidence Interval, CI) and ≤19% since 2017 (average 17%; 15%–19% at 95% CI).

**Figure 1.**
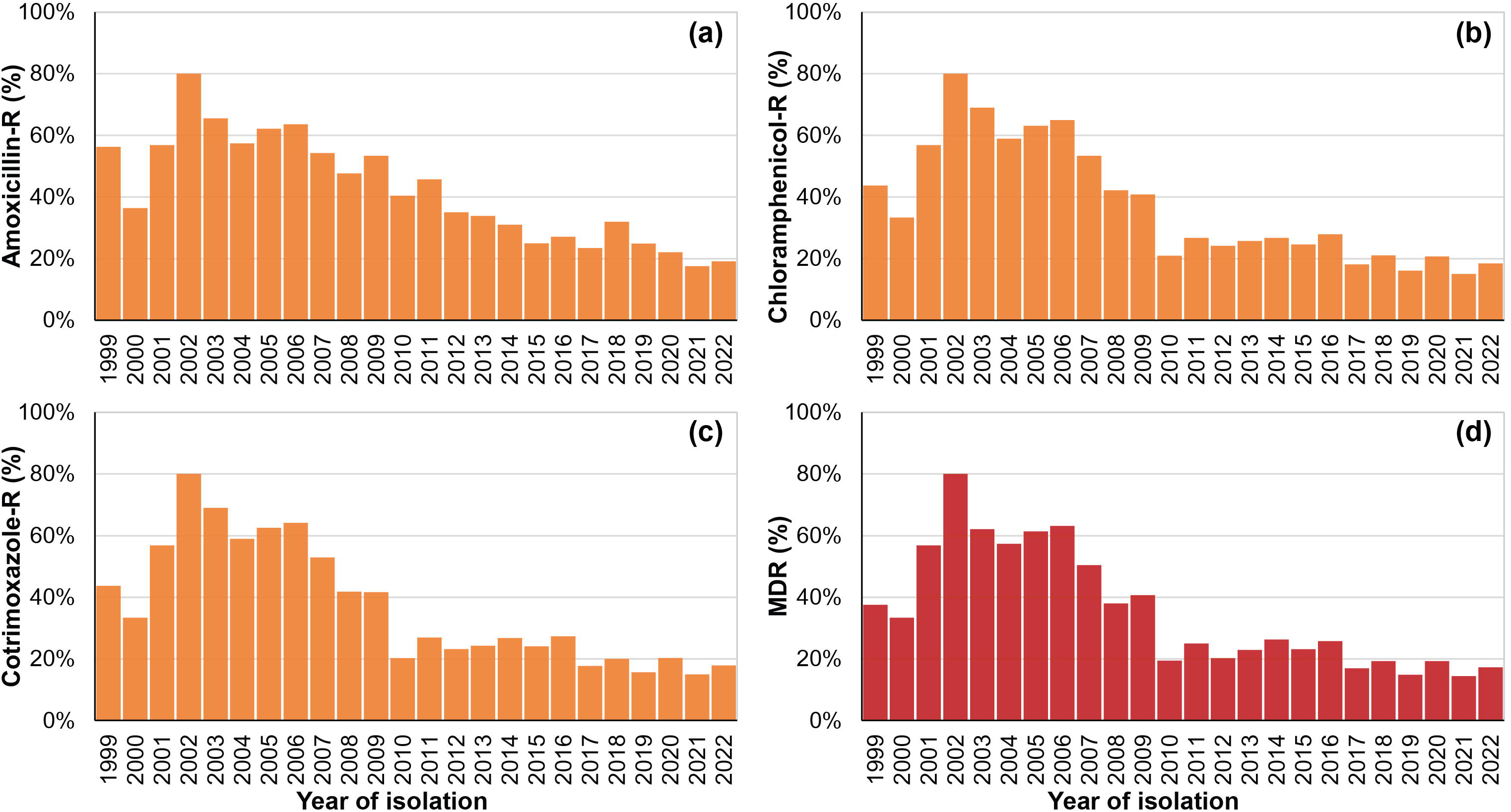
Yearly trends in resistance (-R) and multidrug resistance (MDR) among *Salmonella* Typhi Cases (1999-2022, Bangladesh). (a) Amoxicillin Resistance, (b) Chloramphenicol Resistance, (c) Cotrimoxazole Resistance, and (d) MDR (defined as concurrent resistance to amoxicillin, chloramphenicol, and cotrimoxazole). Information on MDR is presented for all recorded cases.

### Resistance to ciprofloxacin, ceftriaxone, and azithromycin

Non-susceptibility to ciprofloxacin in *Salmonella* Typhi was notably high, reaching 95% in 2001 from 56% in 1999, and remained above 90% throughout the study period. In 2022, 98% (1,266/1,298) of the isolates were non-susceptible to ciprofloxacin (Figure 2a). We further examined the non-susceptibility trend to ciprofloxacin using minimum inhibitory concentration (MIC) data (Figure 3a). Our analysis with the Generalized Additive Model (GAM) of the MIC data revealed a consistent trend with no significant changes in non-susceptibility to ciprofloxacin. The MIC trend for ciprofloxacin remained stable at approximately 0.25 mg/L throughout the entire study period (Figure 3a).

**Figure 2.**
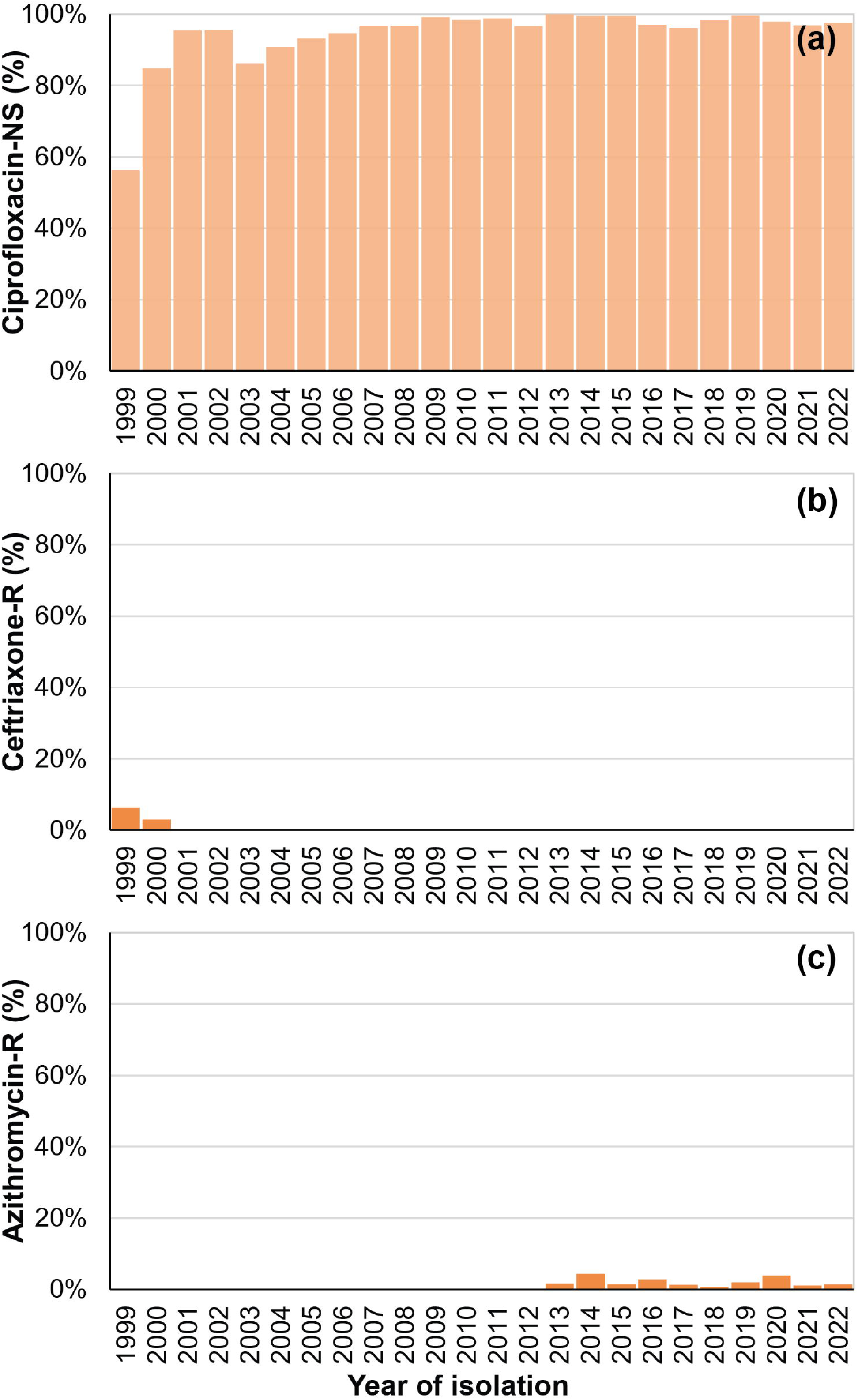
Yearly trends in *Salmonella* Typhi susceptibility for Ciprofloxacin, Ceftriaxone, and Azithromycin (1999-2022) in Bangladesh. **(a)** Ciprofloxacin Non-Susceptibility (NS), **(b)** Ceftriaxone Resistance (-R), and **(c)** Azithromycin Resistance (-R). Susceptibility data were present for 12,431, 12,414, and 8,187 cases respectively.

**Figure 3.**
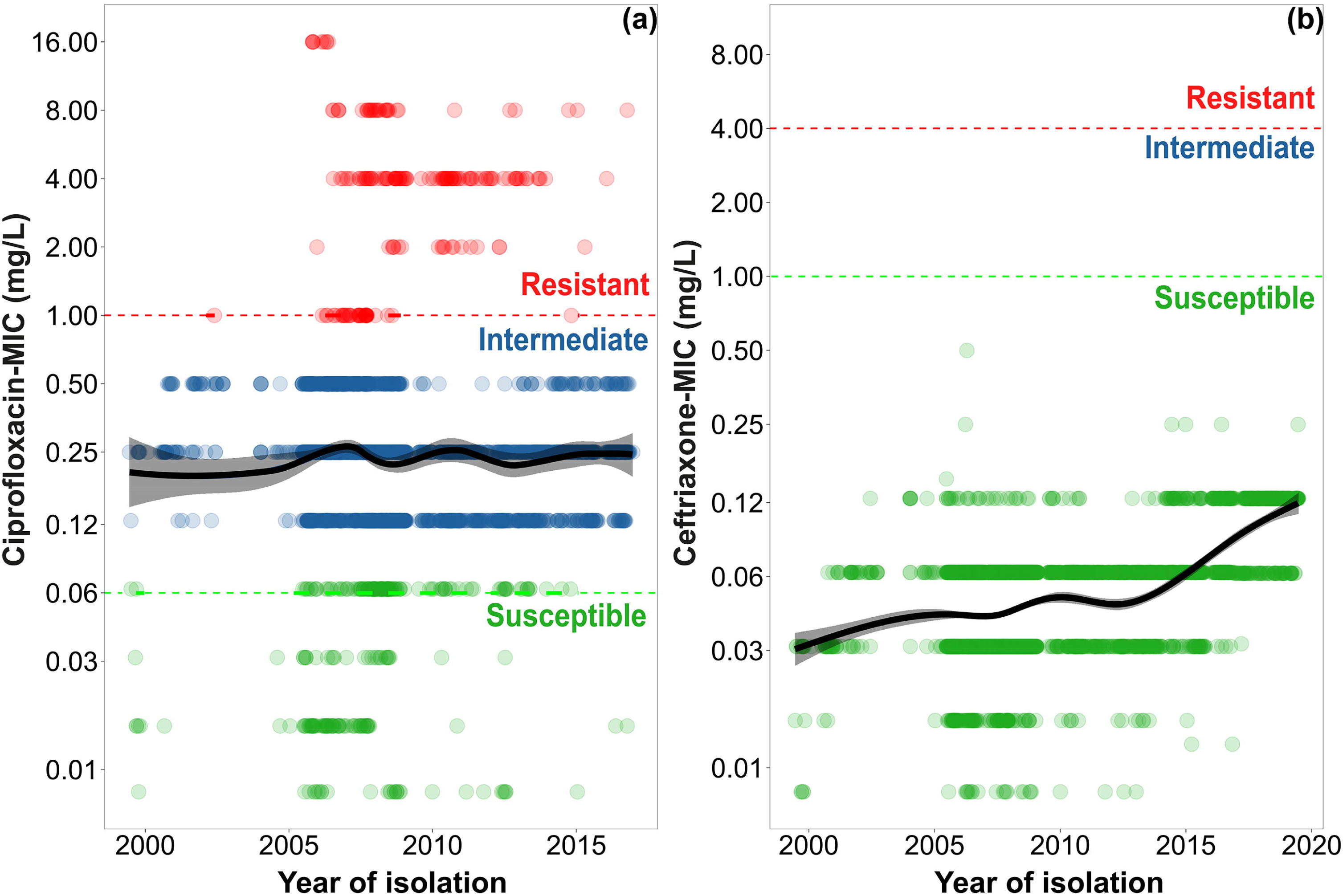
Changes in minimum inhibitory concentrations (MIC) for (a) ciprofloxacin and (b) ceftriaxone among *Salmonella* Typhi cases from 1999 to 2022 in Bangladesh. The MICs were determined for a total of 3,214 and 3,518 isolates for ciprofloxacin (1999–2016) and ceftriaxone (1999–2019), respectively. The color of the dots represents resistant (red), intermediate (blue), and susceptible (green) isolates. The generalized additive model (GAM) lines depict the changes in the MICs (black line), with a 95% confidence interval (grey shades) over time.

Ceftriaxone resistance, on the other hand, was rare among *Salmonella* Typhi isolates during our study period. Only two isolates were ceftriaxone-resistant, one in 1999 and the other in 2000, with no further instances recorded (Figure 2b). To gain detailed insights into this resistance trend, we generated ceftriaxone MIC data for a subset of isolates (n = 3,518) spanning from 1999 to 2019. Our analysis showed a gradual increase in MIC values over time. Overall resistance to ceftriaxone rose from 0.03 mg/L in 2001 to 0.12 mg/L in 2019 (Figure 3b). While these values remained below the breakpoint for reduced susceptisbility (1.0 mg/L as per CLSI standards, red dashed line in Figure 3b), they represent a four-fold increase. A similar fourfold increase was observed in the ceftriaxone MIC_50_ during this period.

Azithromycin resistance in *Salmonella* Typhi was first identified in 2013 during our study period (Figure 2c). Subsequently, an average of 2% (1%–3%; 95% CI) of *Salmonella* Typhi isolates per year displayed resistance to azithromycin. An in-depth analysis using local polynomial regression to examine the yearly percentage of azithromycin-resistant isolates did not show a substantial increase in azithromycin resistance since its initial identification in 2013 (no data shown).

### Correlation between AMR and antimicrobial consumption

To gain deeper insights, we compared AMR data for cotrimoxazole, ciprofloxacin, and azithromycin with national antimicrobial consumption data (Figure 4). Cotrimoxazole consumption exhibited a notable decline, from 0.8 defined daily doses (DDD) per 1,000 persons per day in 1999 to 0.1 DDD/1,000 persons/day in 2020 (Figure 4a). Concurrently, resistance to cotrimoxazole showed a decreasing trend during this period (Figure 1c and 4a). Also, it showed significant correlation with consumption patterns (r = 0.77; p = 0.00 at a 95% confidence interval, CI).

**Figure 4.**
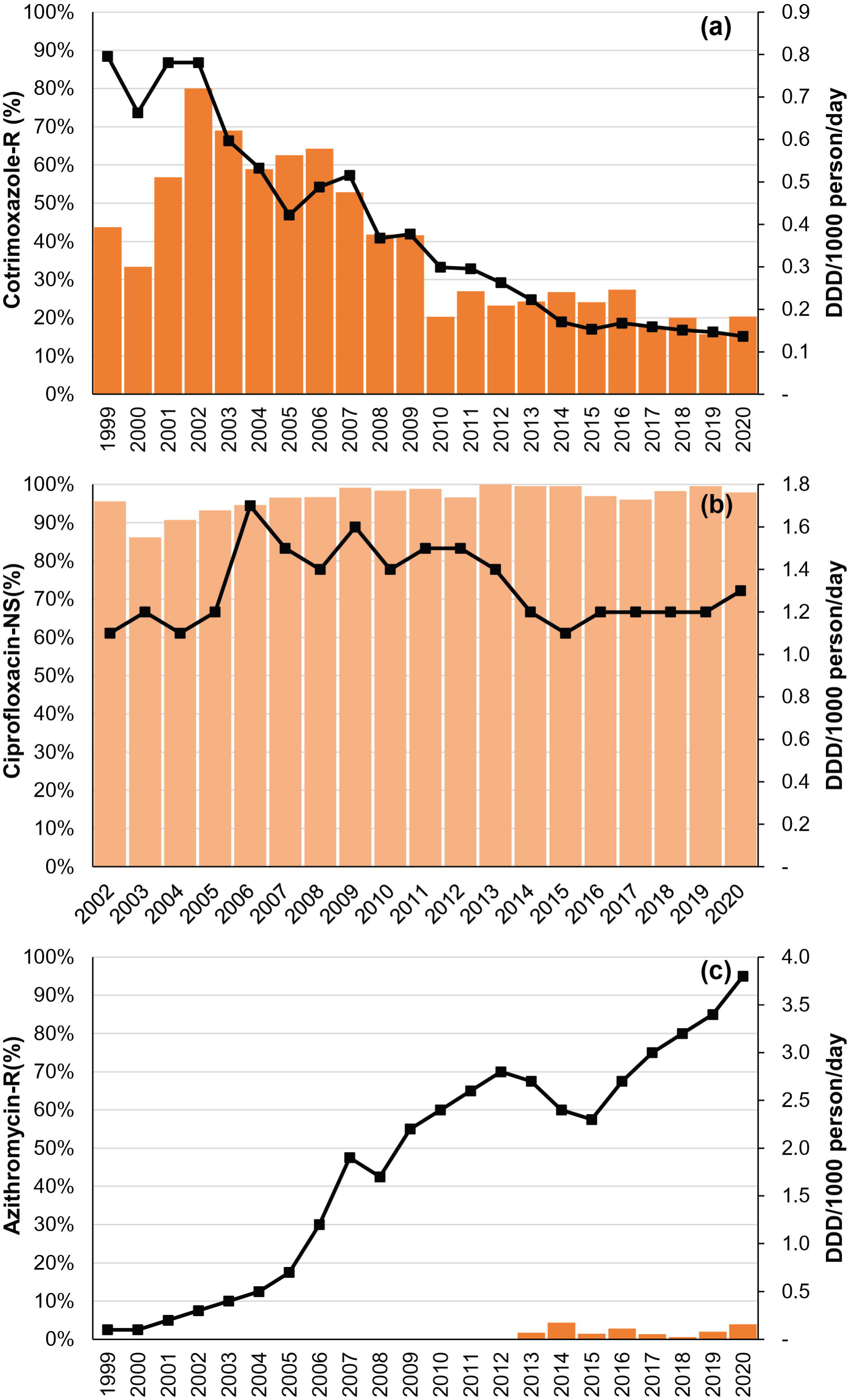
Patterns of antibiotic consumption and antimicrobial resistance in **(a)** Cotrimoxazole, **(b)** Ciprofloxacin, and **(c)** Azithromycin among *Salmonella* Typhi Cases in Bangladesh from 1999 to 2020. Consumption data for ciprofloxacin was unavailable from 1999 to 2001. Resistance (-R) or Non-susceptibility (NS) to cotrimoxazole, ciprofloxacin, and azithromycin are presented on the primary Y-axis. The secondary Y-axis displays antibiotic consumption, represented as the Defined Daily Dose (DDD) per 1,000 persons per day (DDD/1,000 day/year).

Ciprofloxacin consumption remained consistent throughout the study period, between 1.1 and 1.6 DDD/1,000 persons/day (Figure 4b). Non-susceptibility to ciprofloxacin showed a stable pattern as well, but no significant correlation with consumption was identified (r = 0.21; p = 0.38 at a 95% CI). On the other hand, azithromycin consumption increased by 38-fold during study years, 1999-2020, from 0.1 DDD/1,000 persons/day to 3.8 DDD/1,000 persons/day. The average annual increase in azithromycin consumption was 0.18 DDD/1,000 persons/day (0.07–0.29 at a 95% CI) (Figure 4c).

## Discussion

This study provides a comprehensive analysis of 12,435 cases of typhoid fever cases spanning 24 years in Bangladesh, shedding light on AMR patterns of the disease. In endemic countries like Bangladesh, typhoid fever is largely treated in the community settings while the hospitals deal complicated cases. This unique care-seeking behavior made it challenging to estimate the true typhoid burden and extent of AMR.^10, 11^ In addition, hospital-based surveillance from Bangladesh reported higher proportion of MDR typhoid cases than community settings,^10^ potentially not reflecting the true scenario. To bridge this gap, we established a typhoid surveillance system in Dhaka, Bangladesh, covering both hospital and community sites.^5, 8, 32, 33^ Popular Diagnostic Center (PDC), the community-based site and OPD of the two study hospitals contributed 72% (8,957/12,435) of the cases in our study.

We analyzed the dataset to observe trends in AMR among *Salmonella* Typhi isolates over time. Notably, there were declining resistance patterns for first-line antimicrobials, including amoxicillin, chloramphenicol, and cotrimoxazole (Figure 1a–c). This decline was associated with a decrease in multidrug-resistant (MDR) typhoid cases (Figure 1d). Since 2017, over 80% of *Salmonella* Typhi isolates have remained susceptible to first-line drugs, with the average MDR rate during 2017–2022 being 17% (15%–19%, 95% CI). Similar reductions in MDR have been documented in enteric fever surveillances across various countries, including India, Vietnam, Laos, Indonesia, Nepal, and Pakistan (pre-2016 XDR outbreak).^34–41^ The Global Typhoid Genomics Consortium’s report further supports decreasing MDR trends in several typhoid-endemic countries including Bangladesh, India, Nepal, Indonesia, and the Philippines.^42^ Additionally, our data revealed a decreasing trend in cotrimoxazole consumption, a key first-line antimicrobial in Bangladesh (Figure 4a). It aligned with declining resistance to cotrimoxazole over time and demonstrated a significant correlation (r = 0.77; p = 0.00 at 95% CI; Figure 1c and 4a). Further investigation is needed to understand the intricate relationship between drug consumption and resistance.

Ciprofloxacin non-susceptibility and consumption, on the other hand, remained unchanged (Figure 2a and 4b). The ciprofloxacin MIC also revealed continuing non-susceptibility (Figure 3a), indicating its ongoing ineffectiveness for treating typhoid in Bangladesh. The current empirical typhoid treatment in the country is either ceftriaxone or oral azithromycin. Only two ceftriaxone-resistant *Salmonella* Typhi isolates were identified in our study (in 1999 and 2000). However, there was a gradual increase in ceftriaxone MIC from 0.03 to 0.12 mg/L over 20 years (1999-2019; Figure 3b). Although still below the non-susceptibility breakpoint (1.0 mg/L as per the CLSI^43^), the ceftriaxone MIC may approach reduced susceptibility in the near future.

Azithromycin resistance in *Salmonella* Typhi was first identified in our study in 2013, but remained sporadic with only 1% isolates in 2022 (Figure 2c). The consumption of the drug, on the other hand increased by ∼40-fold during 1999-2020 (Figure 4c). High azithromycin use may exert selection pressure, potentially fostering both macrolide- and non-macrolide resistance determinants in the gut.^44^ The underlying molecular cause of azithromycin resistance in *Salmonella* Typhi is a single-point mutation in the AcrB efflux pump gene, *acrB*-717, reported globally.^16, 42, 45^ This mutation also independently emerged in diverse *Salmonella* Typhi genotypes.^45^ Statistical modeling of a random *Salmonella* Typhi genome dataset from Bangladesh suggests a rising effective population size of azithromycin-resistant isolates.^46^ Our study findings do not demonstrate a similar trend (Figure 4c).

The introduction of the typhoid conjugate vaccine (TCV) in Bangladesh has the potential to markedly reduce typhoid burden, including drug-resistant cases. Once introduced, the expected decreasing trend in the post-TCV period will facilitate the focused effort on effective antimicrobial stewardship strategies for Bangladesh. Previous studies showed that for each culture-confirmed typhoid case, at least three patients receive antibiotics in Bangladesh.^47^ The TCV, with its decreasing trend, could significantly cut the antibiotic consumption in the country which is historically high, even increasing according to our results (Figure 4b and 4c).

Before MDR typhoid spread across the globe, the first-line antimicrobials were widely used to treat typhoid. These low-cost antimicrobial agents are well-tolerated and saved millions of lives from typhoid fever for decades (1950s–1980s).^48–50^ Even countries without MDR cases, like Samoa island, continue using amoxicillin as the primary typhoid treatment,^51, 52^ emphasizing the efficacy of these low-cost agents. The successful use of cotrimoxazole to treat typhoid in recent times, has also been reported.^53^ All three first-line drugs have been listed as essential drugs by the Government of Bangladesh since 1982, preventing pharmaceutical price hikes.^54^ Unlike the lengthy and expensive development of new antimicrobials, reintroducing these first-line drugs for treating typhoid cases, requires no research investment. Instead, such step will provide a crucial window for developing new classes of antimicrobial drugs, a process hindered by significant investment barriers in the last decade.^55^

However, reintroducing older drugs may raise concerns about the return of MDR with increased use. The initial H58 *Salmonella* Typhi isolates in the 1990s carried MDR genes on an IncHI1 plasmid. Recent H58 *Salmonella* Typhi isolates in Bangladesh and other endemic countries have integrated these MDR genes into a chromosomal island. This integration helps avoid plasmid-imposed fitness costs, which are negative impacts on bacterial reproduction and survival due to plasmid carriage.^46, 56, 57^ Due to these genomic changes, the re-emergence of MDR would require this H58 lineage to become almost extinct (or at least <5%) before reconsideration. Continuous clinical, laboratory, and genomic surveillance during post-TCV introduction is essential to secure a safe window for the use of the first-line of antimicrobials.^42^ Monitoring antimicrobial use and resistance remains crucial for successfully treating typhoid and other infections effectively.

The results of our study should be considered within the context of several limitations. First, the two hospitals participating in the surveillance were among the largest pediatric reference hospitals in Bangladesh, resulting in a bias towards younger patients. Second, all study sites were in Dhaka, which may not represent other parts of Bangladesh. However, our data reflect the high typhoid burden in the Dhaka city with a reported 57% seropositivity for typhoid fever.^6^ Third, many typhoid patients in Bangladesh are empirically treated without blood culture confirmation. No community-based enteric fever surveillance account for this, limiting comparisons. Finally, national antibiotic consumption data from IQVIA-MIDAS® database were used in this study, as regional or disease-specific data were unavailable.

Taken together, our study describes the trends in antimicrobial resistance (AMR) amongst *Salmonella* Typhi isolates in Bangladesh over the last two decades, correlating them with antibiotic consumption. We proposed a potential antibiotic stewardship strategy, reintroducing first-line antimicrobials for treating typhoid fever. Given their >80% susceptibility for all *Salmonella* Typhi isolates since 2017, this strategy could offer a viable empirical treatment option. Our analysis could serve as a crucial baseline for monitoring the impact of future interventions, including the TCV and improvements in water, sanitation, and hygiene (WASH), on typhoid burden and AMR in the country. The upcoming years are pivotal in combatting typhoid fever, and our study will be instrumental in measuring the effectiveness of interventions aimed at reducing the burden and AMR of this often-neglected tropical disease.

## Acknowledgment

We would like to thank all past and present members of the clinical microbiology and epidemiology team of the Child Health Research Foundation.

## Ethics statement

All study protocols received approval from the Ethics Review Committees (ERC) of Bangladesh Shishu Hospital and Institute (BSHI). Informed written consent and clinical information were obtained from parents/legal guardians for hospitalized cases. For out-patient cases, no formal consent was obtained as blood samples were collected as part of routine clinical care at the discretion of the treating physician and data from routine clinical care were retrospectively included without identifiable information.

## Transparency declarations

All authors declare no conflict of interest.

## Data Availability Statement

All relevant study data are provided in the Supplementary Data S1 of the article.

## Supplementary file legends

**Figure S1.** Overview of the *Salmonella* Typhi database of 12,435 cases collected from 1999– 2022 and MDR percentage in Bangladesh. Yearly hospitalized (IPD) and outpatient (OPD) case numbers (on the left *y*-axis) are presented by the year of isolation. The *x*-axis labels indicate the number of isolates collected each year. Percentages of MDR for IPD and OPD cases per year represented as lines on the right *y*-axis.

**Data S1.** Antimicrobial resistance dataset of 12,435 typhoid cases in Bangladesh from 1999 to 2022 (S=Susceptible, I=Intermediate, and R=Resistant).

