## Supplementary figures and images for "Trends in antimicrobial resistance amongst *Salmonella* Typhi in Bangladesh: a 24-year retrospective observational study (1999–2022)"

### Supplementary Figure S1

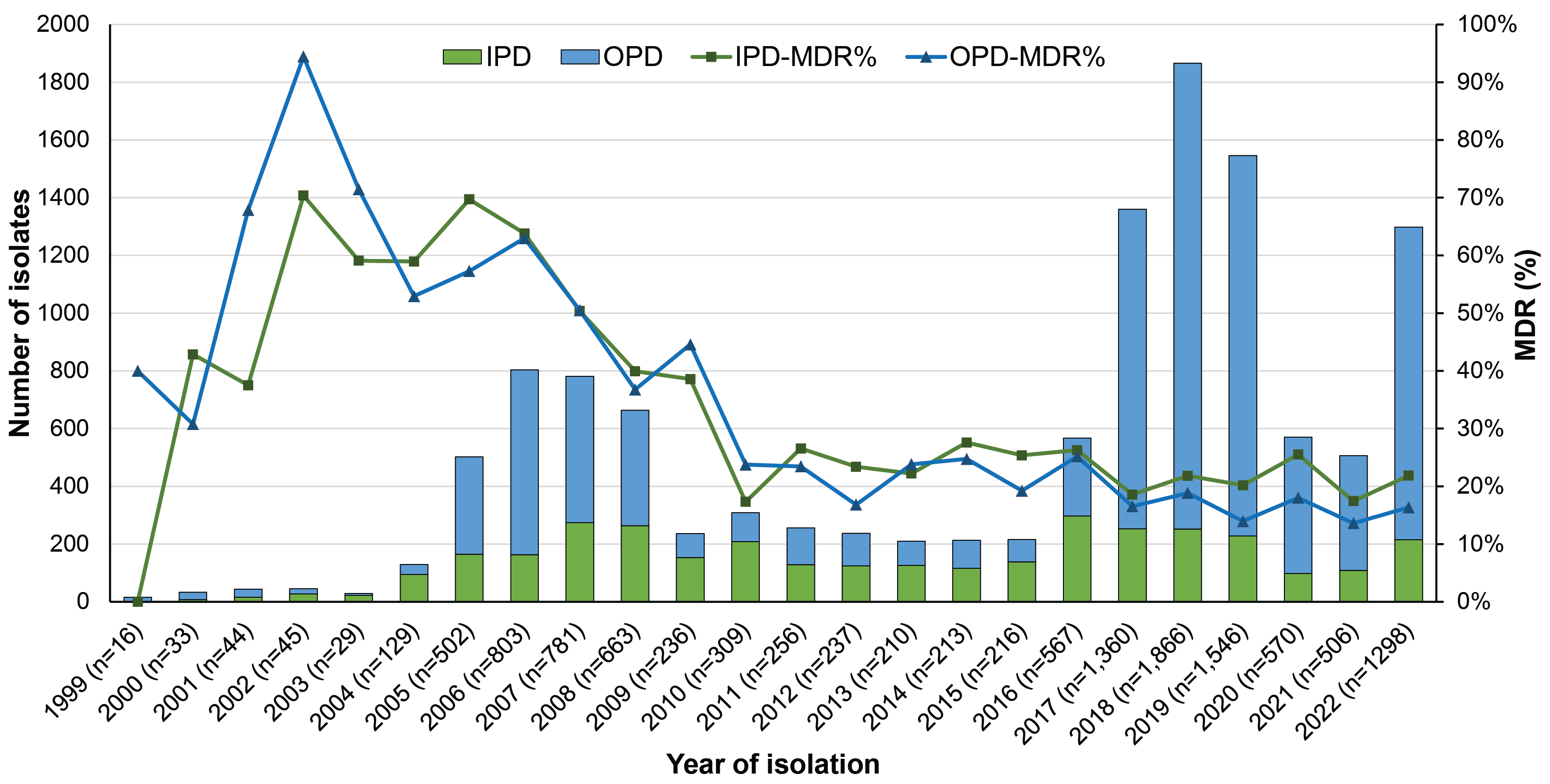
